# BCG vaccination and socioeconomic variables vs Covid-19 global features: clearing up a controversial issue

**DOI:** 10.1101/2020.05.20.20107755

**Authors:** Luigi Ventura, Matteo Vitali, Vincenzo Romano Spica

## Abstract

**Background:** The Covid-19 pandemic is characterized by extreme variability in the outcome distribution and mortality rates across different countries. Some recent studies suggested an inverse correlation with BCG vaccination at population level, while others denied this hypothesis. In order to address this controversial issue, we performed a strict epidemiological study collecting data available on a global scale, considering additional variables such as cultural-political factors and adherence to other vaccination coverages.

**Methods:** Data on 121 countries, accounting for about 99% of Covid-19 cases and deaths globally, were from John’s Hopkins Coronavirus Resource Center, World Bank, International Monetary Fund, United Nations, Human Freedom Report, and BCG Atlas. Statistical models used were Ordinary Least Squares, Tobit and Fractional Probit, implemented on Stata/MP16 software.

**Results:** Based on our results, countries where BCG vaccination is or has been mandated in the last decades have seen a drastic reduction in Covid-19 diffusion (−80% on average) and mortality (−50% on average), even controlling for relative wealth of countries and their governmental health expenditure. A significant contribution to this reduction (respectively −50% and −13% on average) was also associated to the outbreak onset during summer, suggesting a possible influence of seasonality. Other variables turned out to be associated, though to a lesser extent.

**Conclusions:** Relying on a very large dataset and a wide array of control variables, our study confirms a strong and robust association between Covid-19 diffusion and mortality with BCG vaccination and a set socio-economic factors, opening new perspectives for clinical speculations and public health policies.

## Introduction

Since the novel coronavirus SARS-CoV-2 was initially detected in Wuhan, China, in December 2019 (1) more than two million cases of Covid-19 have been confirmed worldwide with a death toll about 140,000 by April 2020, as reported by WHO (https://www.who.int/docs/default-source/coronaviruse/situation-reports/20200417-sitrep-88-covid-191b6cccd94f8b4f219377bff55719a6ed.pdf?sfvrsn=ebe78315_6). One of the puzzles associated to the outspread of this pandemic is the extreme geographic (2) and ethnical (3) variability of its outcomes, both in terms of contagion and mortality with inevitable economic implications (4).

We have witnessed an increased variance in fatality rates as more countries were hit by the virus, generating a clustering of countries in terms of incidence and mortality rates (MR), both across and within affected continents. In Europe, for example, the case fatality rate (CFR) is below or around 3%, such as Portugal, Ireland, Norway and Finland, respectively with 3.45%, 3.79%, 2.35%, 2.35%, but much higher with rates hovering around and above 10% in countries such as Italy, Spain, UK, Sweden, with their respective rates of 13.19%, 10.48%, 13.41%, 10.59% (5).

Especially striking is the contiguity of countries (such as Portugal and Spain, Ireland and UK, Norway/Finland and Sweden) where low and high rates are respectively recorded. Moreover, eastern and south-eastern European countries tend to belong to the first group of countries, those with low incidence and mortality, and south and continental Europe countries to the latter.

By and large, low- and middle-income countries in other continents feature lower fatality and mortality rates, and lower incidence of the epidemic. East Asian countries are characterized, on average, by lower incidence and CFR, with an analogous variability in MRs, that results even higher than the variability in CFR’s (5). It would thus be quite important to understand the main drivers of that variability.

A controversial issue concerns the possible protective role of the Bacillus Calmette–Guérin (BCG) vaccination, opening a still unresolved debate regarding its impact on Covid-19 distribution in countries with different BCG vaccination policies (6-11).

The protective effect of BCG on viral infections had been already suggested by experimental trials, clinical reports and epidemiological studies, and the biological mechanism is supported by different hypothesis, that basically lie in the enhancement of the innate immune response thru trained-immunity processes (12, 13).

The aim of this paper is to robustly investigate the role of BCG vaccination on global incidence, CFRs and MRs of Covid-19 by using linear and nonlinear statistical methods. In so doing, we account for demographic, socioeconomic and health policy confounding factors which, if not properly controlled for, might seriously impair the validity of results and the implications for the following immunological and clinical studies.

## Methods

### Data Source, Sample Selection and Variables

All data are as of April 17^th^, 2020. Data on confirmed cases and deaths come from the John’s Hopkins Coronavirus Resource Center (5). Data on population, geography, income and expenditures, and B Hepatitis, Measles and Diphtheria/Pertussis/Tetanus (DPT) vaccines come from World Development Indicators database (https://databank.worldbank.org/source/world-development-indicators) and from the United Nations Comtrade statistics (https://comtrade.un.org/). Data about foreign direct investments to and from China come from the International Monetary Fund Coordinated Direct Investment Survey (https://data.imf.org/?sk=40313609-F037-48C1-84B1-E1F1CE54D6D5). Detailed information on tuberculosis vaccination policies comes from the Bacille Calmette-Guérin (BCG) vaccine Atlas (14), last updated in 2017 in the online version (http://www.bcgatlas.org/index.php). Data about human freedom comes from the 2019 Human Freedom Report by the Fraser Institute (https://www.cato.org/sites/cato.org/files/human-freedom-index-files/human-freedom-index-2018-revised.pdf).

Data from a total of 121 countries, out of the 209 that reported cases of Covid19, accounting for about 99% of both confirmed cases and deaths, have been used. The countries in the analysis, listed in supplementary appendix, have been chosen in view of the availability of observations relative to covariates.

The set of dependent and independent variables is reported in Table 1. In particular, we used confirmed cases per million inhabitants as a proxy for the intensity of contagion; the number of cases 15 days earlier as a proxy for the stage of the diffusion of the virus; population in the largest city as a proxy for density and the degree of urbanization; life expectancy at birth as a comprehensive health indicator, and as a proxy for the share of aged people in the population; the latitude to define both the season as of April 17^th^ (above or below the Equatorial line) and tropical countries (those countries whose latitude as defined by the corresponding variable in the World Development Indicators lies in between the two tropics). As for BCG vaccination policy, two alternative continuous measures were constructed, and used for robustness checks: the BCG coverage, as reported in national surveys in various years, and the years of absence of mandated vaccination, until 2020.

**Table 1.** Summary statistics for dependent and independent variables

| **Dependent Variables** | **Mean** | **SD<sup>a</sup>** | **N of countries** |
| --- | --- | --- | --- |
| Confirmed cases x million inhabitants (n°) | 469·73 | 791·51 | 121 |
| Case Fatality Rate (%) | 4·10 | 3·99 | 121 |
| Mortality rate (x 1,000 people) | 0·03 | 0·08 | 121 |
| **Independent Variables** |  |  |  |
| Positives for SARS-CoV-2 15 days earlier (n°) | 8,194·84 | 28,885·97 | 121 |
| Population in largest city (n° x 10,000) | 466·52 | 620·42 | 121 |
| Rural population (% of the total population) | 37·73 | 22·42 | 121 |
| Migrant population (% of the total population) | 9·08 | 15·12 | 121 |
| Population (n° x 10,000) | 5,672·12 | 17,681·96 | 121 |
| Life expectancy at birth (years) | 72·63 | 7·83 | 121 |
| GDP<sup>b</sup> per capita (USD<sup>c</sup> thousands) | 19·67 | 19·90 | 121 |
| Domestic private health expenditure per capita (USD<sup>c</sup>) | 470·31 | 454·69 | 121 |
| Domestic general governmental health expenditure (% GDP<sup>b</sup>) | 3·79 | 2·50 | 121 |
| Imports from China (USD<sup>c</sup> millions) | 18,781·65 | 54,388·69 | 121 |
| Foreign Direct Investment to China (USD<sup>c</sup> billions) | 16·50 | 126·58 | 121 |
| Foreign Direct Investment from China (USD<sup>c</sup> billions) | 10·41 | 87·28 | 121 |
| Summertime (yes/no) | 0·18 | 0·39 | 121 |
| Tropical (yes/no) | 0·45 | 0·50 | 121 |
| Landlocked (yes/no) | 0·21 | 0·41 | 121 |
| Human Freedom Index<sup>d</sup> (0 to 10) | 6·93 | 1·04 | 121 |
| B Hepatitis vaccination (% 1 year-old children) | 86·98 | 12·94 | 115 |
| Measles vaccination (% 12-23 months-old children) | 87·87 | 12·70 | 115 |
| DPT<sup>e</sup> vaccination (% 12-23 months-old children) | 87·85 | 12·64 | 115 |
| Compulsory<sup>f</sup> BCG vaccine (yes/no) | 0·95 | 0·22 | 121 |
| BCG coverage (% of the total population) | 0·86 | 0·22 | 51 |
| Absence of BCG vaccination from 2020 backwards (years) | 4·19 | 11·11 | 116 |

<sup>a</sup>Standard Deviation; <sup>b</sup>Gross Domestic Product; <sup>c</sup>United States Dollar; <sup>d</sup>10 denoting the most freedom;
<sup>e</sup>Diphtheria/Pertussis/Tetanus; <sup>f</sup>presently and in the last 45 years

Coverage rates for different vaccines (B Hepatitis, Measles and DPT) were also used, to disambiguate the effects of BCG from those of a more general vaccination policy.

Among the variables proxying for economic ties with China, where the epidemic first appeared, we include imports from China, and the levels of inward and outward Foreign Direct Investment (FDI) relative to China. Finally, to proxy for the compliance with the lockdown measures implemented by the various governments, we use the Index of Human Freedom (HFI), a weighted average of 79 distinct indicators (37 for the personal freedom subindex and 42 for the economic freedom subindex), each one ranging from 0 to 10, with 10 representing the most freedom. The HFI ranges therefore from 0 to 10, in increasing order of freedom (https://www.cato.org/sites/cato.org/files/human-freedom-index-files/human-freedom-index-2018-revised.pdf).

We used Gross Domestic Product (GDP) per capita, and private and general government health expenditure to proxy for countries’ level of development (general and of their health system) and for the countries’ testing capability (more income and a richer health system should be positively correlated to more Covid-19 testing).

### Statistical Analysis

To model our dependent variables, we used both ordinary least squared, as a reference estimator, and nonlinear estimation methods. In particular Tobit regressions, estimating both the impacts of covariates on the probability of a country reporting more than 100 cases as of April 17^th^, and their effect on relative diffusion, was our preferred estimation method.

The reported coefficients in the Tobit regression represent the marginal effects of the explanatory variables on the outcome variable, after accounting for the inclusion of countries in the high incidence group.

The second and third outcome variables, i.e. CFRs and MRs, were first modelled by ordinary least squares to obtain benchmark estimations, and then by Probit fractional regression methods to account for the fractional nature of the dependent variables (15). When the dependent variable is a fraction, as with CFRs and MRs, using log-odds transformation or Tobit regressions with lower and upper limits set to 0 and 1 may yield biased results (15, 16). Therefore, fractional regressions will be our preferred estimation method for fatality and mortality rates.

For consistency and comparison purposes all models included the same set of explanatory variables. Moreover, ordinary least squares and fractional regressions also account for heteroskedasticity, by using robust variance-covariance estimators.

All statistical analyses have been performed by using Stata/MP 16 for Windows.

## Results

Table 2 reports the regression results for relative incidence (reported cases over total population) obtained by OLS, Tobit without controlling for other vaccines and Tobit controlling for other vaccines. For both OLS and Tobit, the estimated coefficients represent the marginal effect of the covariates on the outcome variable. Table 2 shows large and strongly significant effects for per capita gross domestic product and Human Freedom Index (positive) and for the summer season and BCG vaccination (negative), even controlling for more vaccinations.

**Table 2.**
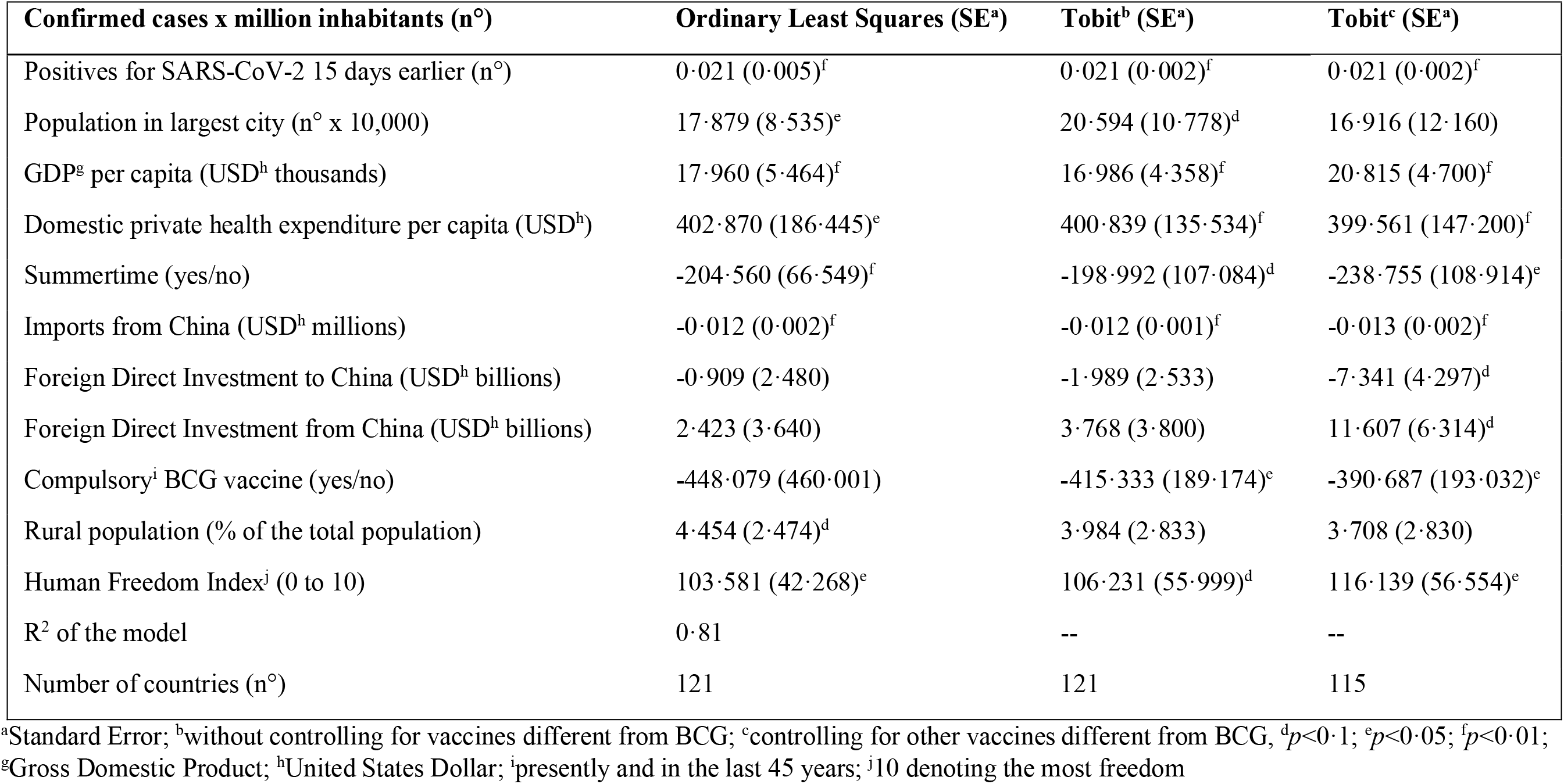
Determinants of the relative number of confirmed cases. OLS and Tobit estimates for the share of confirmed cases over total population as the dependent variable. Tobit model with left censoring at 100 cases as of April 17^th^, and coefficients represent marginal effects on the dependent variable

| Confirmed cases x million inhabitants (n°) | Ordinary Least Squares (SE<sup>a</sup>) | Tobit<sup>b</sup> (SE<sup>a</sup>) | Tobit<sup>c</sup> (SE<sup>a</sup>) |
| --- | --- | --- | --- |
| Positives for SARS-CoV-2 15 days earlier (n°) | 0.021 (0.005)<sup>f</sup> | 0.021 (0.002)<sup>f</sup> | 0.021 (0.002)<sup>f</sup> |
| Population in largest city (n° x 10,000) | 17.879 (8.535)<sup>e</sup> | 20.594 (10.778)<sup>d</sup> | 16.916 (12.160) |
| GDP<sup>g</sup> per capita (USD<sup>h</sup> thousands) | 17.960 (5.464)<sup>f</sup> | 16.986 (4.358)<sup>f</sup> | 20.815 (4.700)<sup>f</sup> |
| Domestic private health expenditure per capita (USD<sup>h</sup>) | 402.870 (186.445)<sup>e</sup> | 400.839 (135.534)<sup>f</sup> | 399.561 (147.200)<sup>f</sup> |
| Summertime (yes/no) | -204.560 (66.549)<sup>f</sup> | -198.992 (107.084)<sup>d</sup> | -238.755 (108.914)<sup>e</sup> |
| Imports from China (USD<sup>h</sup> millions) | -0.012 (0.002)<sup>f</sup> | -0.012 (0.001)<sup>f</sup> | -0.013 (0.002)<sup>f</sup> |
| Foreign Direct Investment to China (USD<sup>h</sup> billions) | -0.909 (2.480) | -1.989 (2.533) | -7.341 (4.297)<sup>d</sup> |
| Foreign Direct Investment from China (USD<sup>h</sup> billions) | 2.423 (3.640) | 3.768 (3.800) | 11.607 (6.314)<sup>d</sup> |
| Compulsory<sup>i</sup> BCG vaccine (yes/no) | -448.079 (460.001) | -415.333 (189.174)<sup>e</sup> | -390.687 (193.032)<sup>e</sup> |
| Rural population (% of the total population) | 4.454 (2.474)<sup>d</sup> | 3.984 (2.833) | 3.708 (2.830) |
| Human Freedom Index<sup>j</sup> (0 to 10) | 103.581 (42.268)<sup>e</sup> | 106.231 (55.999)<sup>d</sup> | 116.139 (56.554)<sup>e</sup> |
| R<sup>2</sup> of the model | 0.81 | -- | -- |
| Number of countries (n°) | 121 | 121 | 115 |

<sup>a</sup>Standard Error; <sup>b</sup>without controlling for vaccines different from BCG; <sup>c</sup>controlling for other vaccines different from BCG, <sup>d</sup> $p < 0.1$ ; <sup>e</sup> $p < 0.05$ ; <sup>f</sup> $p < 0.01$ ;
<sup>g</sup>Gross Domestic Product; <sup>h</sup>United States Dollar; <sup>i</sup>presently and in the last 45 years; <sup>j</sup>10 denoting the most freedom

Table 3 contains estimated coefficients, with corresponding standard errors and marginal effects, for CFR and MRs fractional regressions. This table reveals large and strongly significant effects for health expenditure and tropical position (positive) and for summer season and, above all, BCG vaccination (negative), even controlling for more vaccinations.

**Table 3.**
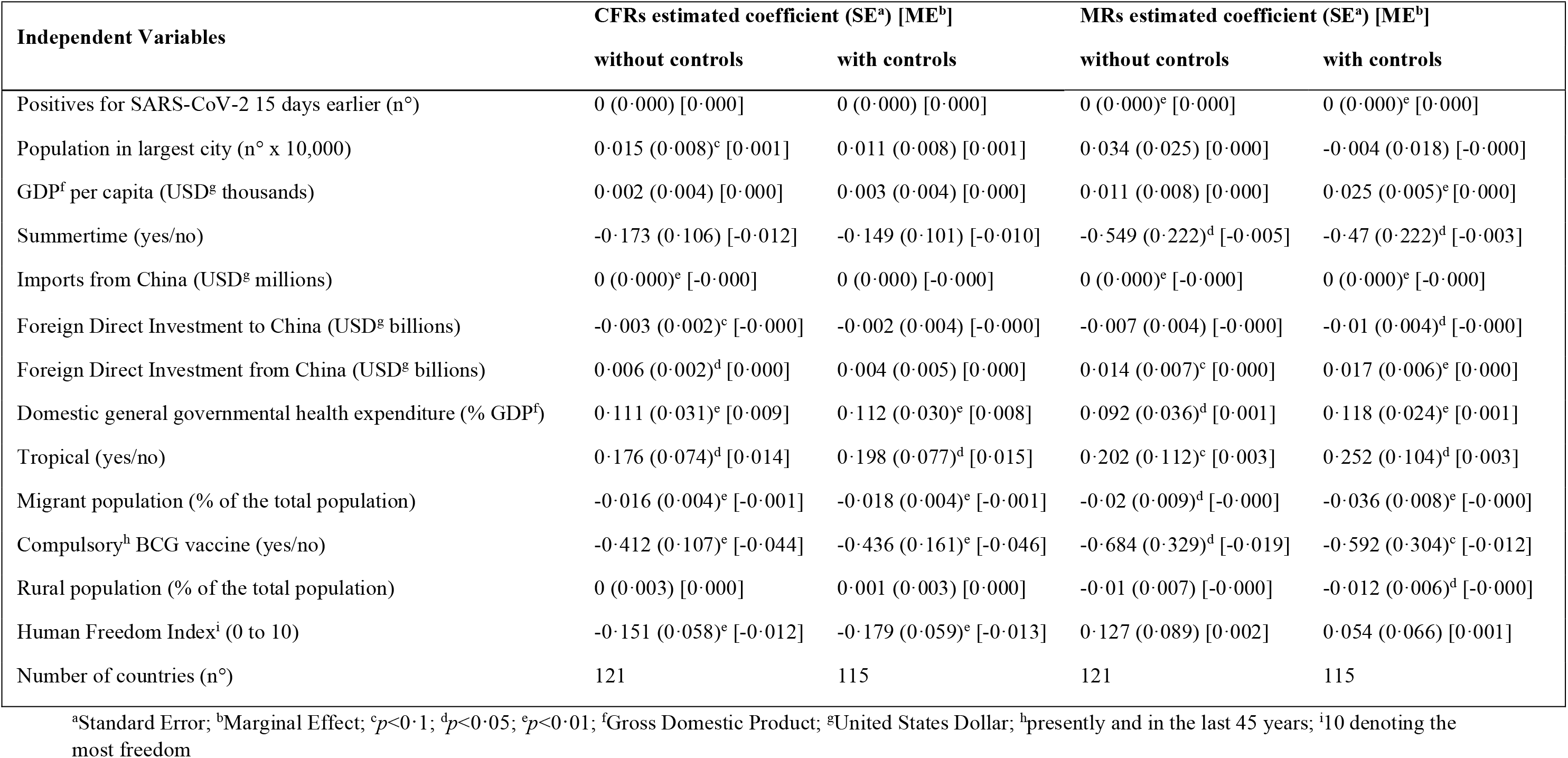
Determinants of Case Fatality Rates (CFRs) and Mortality Rates (MRs). Probit fractional regressions estimates for the sample without and with the additional vaccination controls (B hepatitis, Measles, DTP)

Table 4 contains the results of a robustness analysis on relative incidence, where alternative measures for BCG have been used as explanatory variables instead of the BCG dummy.

**Table 4.** Determinants of Relative Incidence and alternative BCG variables. Tobit estimates for the share of confirmed cases over total population as the dependent variable, with left censoring at 100 cases as of April 17^th^, and coefficients represent marginal effects on the dependent variable

| Independent Variables | Tobit<sup>a</sup> (SE<sup>b</sup>) | Tobit<sup>c</sup> (SE<sup>b</sup>) |
| --- | --- | --- |
| Positives for SARS-CoV-2 15 days earlier (n°) | 0·022 (0·002)<sup>f</sup> | 0·030 (0·009)<sup>f</sup> |
| Life expectancy at birth (years) | 20·640 (11·318)<sup>d</sup> | 23·358 (19·226) |
| GDP<sup>g</sup> per capita (USD<sup>h</sup> thousands) | 21·276 (4·687)<sup>f</sup> | 21·050 (11·930)<sup>d</sup> |
| Domestic private health expenditure per capita (USD<sup>h</sup>) | 306·987 (144·401)<sup>e</sup> | 271·580 (316·930) |
| Summertime (yes/no) | -233·147 (107·458)<sup>e</sup> | 40·292 (170·410) |
| Imports from China (USD<sup>h</sup> millions) | -0·011 (0·002)<sup>f</sup> | -0·002 (0·005) |
| Foreign Direct Investment from China (USD<sup>h</sup> billions) | 10·611 (6·367)<sup>d</sup> | -36·453 (41·706) |
| Migrant population (% of the total population) | -8·568 (4·579)<sup>d</sup> | 15·804 (15·070) |
| Absence of BCG vaccination from 2020 backwards (years) | 8·978 (4·463)<sup>e</sup> | -- |
| Human Freedom Index<sup>i</sup> (0 to 10) | 91·987 (57·711) | 221·048 (116·135)<sup>d</sup> |
| BCG coverage (% of the total population) | -- | -724·321 (329·727)<sup>e</sup> |
| Number of countries (n°) | 115 | 46 |

<sup>a</sup>absence of BCG vaccination from 2020 backwards; <sup>b</sup>Standard Error; <sup>c</sup>BCG coverage <sup>d</sup> $p<0\cdot1$ ; <sup>e</sup> $p<0\cdot05$ ; <sup>f</sup> $p<0\cdot01$ ; <sup>g</sup>Gross Domestic Product; <sup>h</sup>United States Dollar; <sup>i</sup>10 denoting the most freedom

Table 5 contains the results of the same robustness analysis performed on the CFRs and MRs. Both tables 4 and 5 confirm the results obtained with the previous regressions.

**Table 5.**
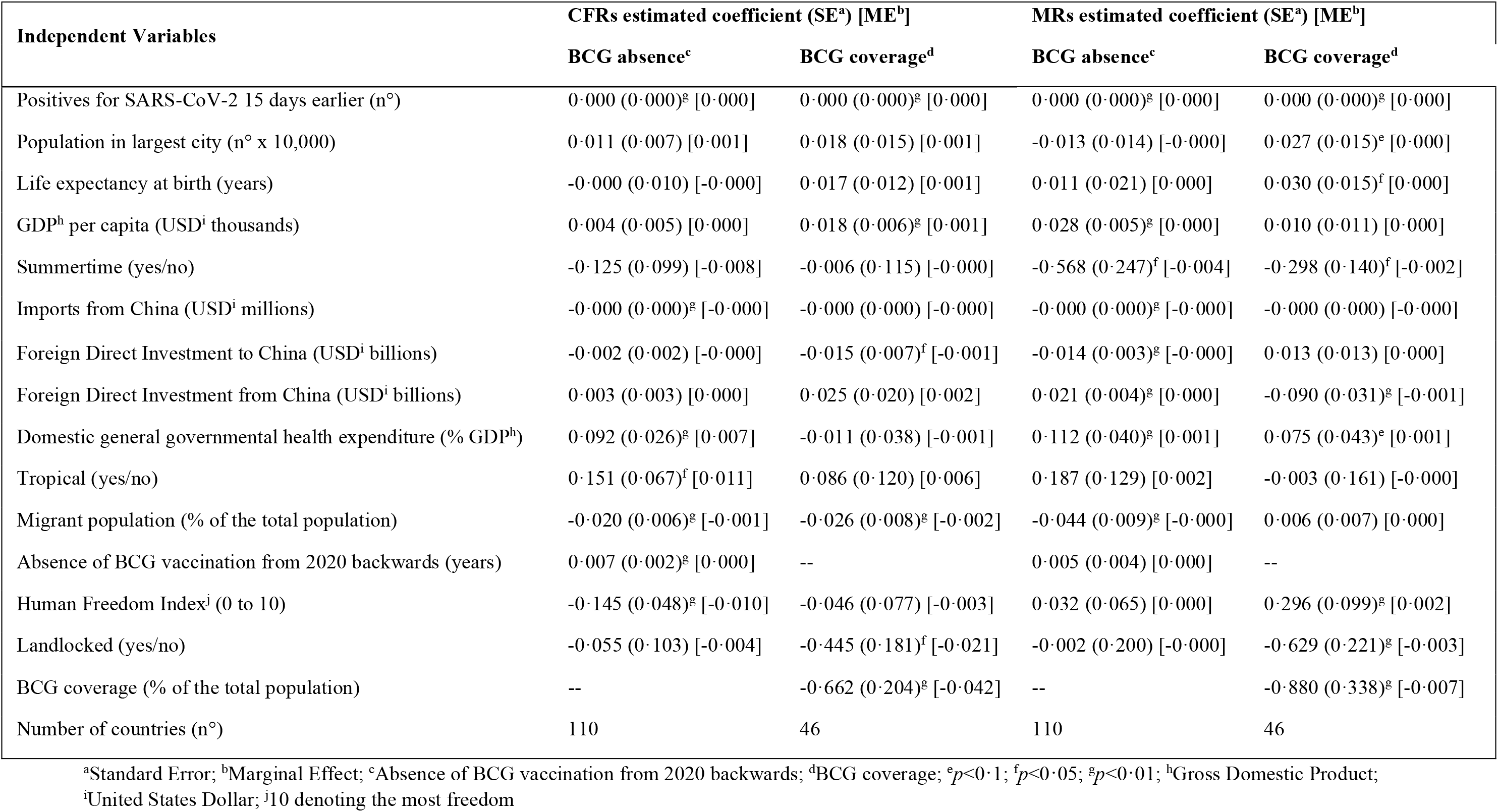
Determinants of CFR and mortality, alternative BCG variables. Probit fractional regression estimates for CFRs and MRs, with alternative measures for BCG vaccination

All regressions feature the same set of explanatory variables, listed in the data section, but tables only report statistically significant coefficients, as well as their respective significance (10%, 5%, or 1%).

## Discussion

To the best of our knowledge, this is the first study assessing the impact of BCG vaccination on the diffusion and mortality of Covid-19 at the global level by controlling for a comprehensive set of social, economic, geographic and demographic variables. This allows to greatly reduce the risk of spurious correlations among variables and confers high statistical robustness to the results which are therefore more amenable to causal interpretation.

From the second and third column in Table 2, containing the statistically significant Tobit coefficients of relative incidence, we observe that the number of positive cases 15 days earlier has a very significant and sizeable positive coefficient, capturing the different stage of the epidemic across countries (a larger number of cases implies a higher probability of contagion).

Among the demographic variables, population in the largest city has a very sizeable, positive and statistically significant coefficient, indicating that high urban density fosters the epidemic. Other demographic variables do not reach statistical significance, possibly in view of high variability in data. Lastly, the percentage of immigrant over total population is negatively correlated with the extent of the epidemic, but with a non statistically significant coefficient. Its sign, however, might be read in the light of the impact of BCG vaccine, as explained below. Or it might be read as resulting from the lower probability of those immigrant communities being tested, as suggested by Borjas (17).

Among geographical variables, the dummy for Summer (countries below Equatorial Line) is negative, large, and highly statistically significant. This strongly reinforces results in Ozdemir and colleagues (7), finding that the mean of cases per population ratio was higher in the Northern hemisphere. The magnitude of the coefficient is really large, and indicates that countries in the Southern hemisphere have, on average and all else equal, over 200 cases per million less (against a mean value of confirmed cases per million inhabitants of about 470). Seasonality is a key question for describing the trend of pandemic and predicting future transmission dynamics (18).

Among the economic and health policy variables, GDP has a large and highly significant coefficient in all specifications, which corresponds to our a priori expectations, in view of the maintained hypothesis of a positive correlation between income and number of tests performed. In other words, including the per capita GDP variable allows to control for different testing policies implemented across countries. Domestic private health expenditure also features a positive coefficient, which can most likely be explained in the same way.

The main and well taken criticism, addressed by Curtis and colleagues (6) and Faust and colleagues (https://naturemicrobiologycommunity.nature.com/users/36050-emily-maclean/posts/64892-universal-bcgvaccination-and-protection-against-covid-19-critique-of-an-ecological-study), to most of the recent ecological studies assessing the impact of BCG vaccination upon Covid-19 related outcomes goes as follows: different testing policies, induced by nation-wide economic situations, might impair the validity of results, by spuriously inducing a negative correlation between BCG vaccination (more frequent in low or middle income countries, that on average have less economic means to carry out a substantial testing policy), reported cases and deaths.

In our study we avoid this problem by explicitly accounting for the relative wealth of countries and of their respective health systems, thus netting out the relationship between income and testing policies, and by using a large dataset including quite a few high income countries where BCG vaccine is also mandatory (Japan, as a notable example).

Closer economic ties to China exert an ambiguous effect. On the one hand, imports from China have a negative and statistically significant effect on the extent of the epidemic, most likely because countries closer to China were affected to a limited extent. On the other, FDI from China have a positive, but limited effect, and might derive from the more frequent personal contacts between Chinese and western businessmen, around the start of the crisis. Anyway, if ties with China might have explained the diffusion of the epidemic at earlier stages, they no longer seem to do so.

The HFI has a strong and significant impact on the incidence of the epidemic, suggesting that in freer countries lockdown measures were milder, and that compliance might have been lower in such countries.

Most importantly, the BCG dummy variable has a strong and negative impact in the Tobit specifications. This finding corroborates in a more comprehensive and robust framework those presented in some recent contributions (7-10) and arguing in favor of a correlation between universal BCG vaccination policy and reduced morbidity and mortality for COVID-19. Another study by Dayal and Gupta (11) obtains similar conclusions, comparing CFR’s of countries where BCG re-vaccination is adopted vs. those countries where vaccination is practiced only once in lifetime.

As the significance of BCG might be spurious, driven by the correlation with other vaccinations, the third column in Table 2 contains a second Tobit specification, where the adoption of other vaccination policies (namely, B Hepatitis, Measles and DPT) is also controlled for. Results are substantially unaltered, hinting at a very robust effect of BCG vaccination in reducing the (symptomatic) diffusion of the epidemic.

Tables 3 contains results for CFR’s and MRs in the fractional model specifications. The second and fourth column include two additional vaccination variables (B Hepatitis has not been omitted here in view of its high correlation with DPT) to account for the potential endogeneity of the BCG variable, in view of the possibility of omitted vaccination variables.

Looking at the estimated coefficients, we notice that general government health expenditure, as a percentage of GDP, has a positive and significant effect, possibly revealing a more accurate control of deaths, while closer ties with China have once again ambiguous effects. Negative for imports and for FDI to China, and positive for FDI from China, most likely related to the relative stance of Asian and Western countries in those areas. The impact of all those variables, however, is quite limited in size (less than 1%).

Population wise, while concentration in largest city exerts, not surprisingly, a positive effect on CFRs and MRs (but only significant for CFRs), the percentage of migrant population has a significant and negative effect. For example, one standard deviation of migrant population share would reduce the CFR by about 1. 5%. This might be interpreted in two different ways. On the one hand, paralleling the arguments in Borjas (17), immigrants’ communities might be less checked, and report less fatalities. However, migrants to western countries usually come from countries where BCG is mandatory, which might result associated with lower MRs. More disaggregated data - possibly at individual level - may provide additional information on vulnerability drivers involved in Covid-19 related outcomes (19).

In fact, the most important effect on CFRs and MRs, as for relative incidence, is exerted by the BCG variable, which is associated to a strongly significant reduction of CFR and MR by, respectively, −4.5 percentage points (both with and without additional vaccination controls) and - 1.9 (−1.2 with additional vaccination controls) percentage points. To get a relative idea of the magnitude of these estimates, let us just notice that the mean values of CFRs and MRs in the estimation sample are, respectively, 4.1% and 2.8%.

Anecdotal evidence, especially in Europe, seems largely consistent with the large explanatory power of BCG on CFRs and MRs. We mentioned the wide and puzzling differences in CFRs and MRs between contiguous countries, such as Portugal and Spain, Ireland and UK, Norway/Finland and Sweden. In all those pairs of countries, the first has mandatory BCG vaccination or had it until recent times, the second has not.

Last but not least, the HFI turns out to have a negative and significant effect on the CFR. This, however, might only be due to the results reported above, i.e. the positive effect of HFI on the number of reported cases, which is the denominator of CFR.

As a further control of the robustness of these results, more checks have been performed by replacing the BCG dummy with two different continuous variables related to this vaccination policy. One is BCG coverage, as reported in national surveys. Another is the number of years of missing vaccination, until 2020. Unfortunately, data on coverage were available for a more limited number of countries, but even so the results seem noteworthy. Table 4 contains the results of two Tobit regressions of the relative number of cases, by using the alternative BCG variables. Table 5 reports the results of fractional regressions on both CFRs and MRs, using the two alternative BCG variables. Tables 4 and 5 show a strong and negative impact, of magnitude comparable to that obtained for the BCG variable in the previous analyses, for the BCG coverage variable. Missing years of vaccination also feature a coefficient with the expected positive sign, but only significant in the case of relative incidence and CFRs.

## Conclusions

Our study, by integrating a wide set of controls, attenuates if not eliminates altogether the effects of possible confounding factors affecting previous studies on BCG and Covid-19. The nonlinear, probabilistic methods used in the analyses confer additional statistical robustness to the results. This way, we are able to confirm a robust and large effect of BCG vaccination on Covid-19 diffusion and mortality, and to uncover other noteworthy and potentially relevant statistical relationships. In this wider context, the association between BCG vaccination and Covid-19 diffusion and mortality is fully addressed by a comprehensive epidemiological perspective, that can support novel hypothesis as well as clinical or experimental studies in the field.

## Data Availability

All data sources are open and are all reported in the manuscript.

## Supplementary data

Supplementary data are available at IJE online.

## Acknowledgements

We deeply thank Charles Yuji Horioka and Yoko Niimi for discussions, key insights, comments, and crucial information about the main topic of this paper. Useful advice and help from Maria Ventura and Lory Marika Margarucci is also gratefully acknowledged.

## Author Contributions

All authors contributed equally, interpreted the findings, contributed to writing the manuscript, and approved the final version for publication.

## REFERENCES

1. Zhu N, Zhang D, Wang W, Xingwang L, Bo Y, Jingdong S, Xiang Z, Baoying H, Weifeng S, Roujian L, Peihua N, Faxian Z. 2020. A novel coronavirus from patients with pneumonia in China, 2019. N Engl J Med 382:727–33. https://doi.org/10.1056/NEJMoa2001017.

2. Khan S, Siddique R, Shereen MA, Ali A, Liu J, Bai Q, Bashir N, Xue M. 2020. Emergence of a Novel Coronavirus, Severe Acute Respiratory Syndrome Coronavirus 2: Biology and Therapeutic Options. J Clin Microb 58: e00187–20. https://doi.org/10.1128/JCM.00187-20.

3. Chowkwanyun M, Reed AL. 6 May 2020. Racial health disparities and Covid-19 - Caution and Context. N Engl J Med. https://doi.org/10.1056/NEJMp2012910.

4. Wang Z, Tang K. 2020. Combating COVID-19: health equity matters. Nat Med 26:458. https://doi.org/10.1038/s41591-020-0823-6.

5. Dong E, Du H, Gardner L. 2020. An interactive web-based dashboard to track COVID-19 in real time. Lancet Infect Dis 20:533–4. https://doi.org/10.1016/S1473-3099(20)30120-1.

6. Curtis N, Sparrow A, Ghebreyesus TA, Netea MG. 30 April 2020. Considering BCG vaccination to reduce the impact of COVID-19. Lancet. https://doi.org/10.1016/S0140-6736(20)31025-4.

7. Ozdemir C, Kucuksezer UC, Tamay ZU. 24 April 2020. Is BCG vaccination effecting the spread and severity of COVID-19? Allergy. https://doi.org/10.1111/all.14344.

8. Miller A, Reandelar MJ, Fasciglione K, Roumenova V, Li Yan, Otazu GH. 28 March 2020. Correlation between universal BCG vaccination policy and reduced morbidity and mortality for COVID-19: an epidemiological study. medRxiv https://doi.org/10.1101/2020.03.24.20042937.

9. Gursel M, Gursel I. 6 April 2020. Is global BCG vaccination coverage relevant to the progression of SARS-CoV-2 pandemic? Med Hypotheses. https://doi.org/10.1016/j.mehy.2020.109707.

10. Redelman-Sidi G. 27 April 2020. Could BCG be used to protect against COVID-19? Nat Rev Urol. https://doi.org/10.1038/s41585-020-0325-9.

11. Dayal D, Gupta S. 19 April 2020. Connecting BCG Vaccination and COVID-19: Additional Data. medRxiv https://doi.org/10.1101/2020.04.07.20053272.

12. Moorlag SJCFM, Arts RJW, van Crevel R, Netea MG. 2019. Non-specific effects of BCG vaccine on viral infections. Clin Microbiol Infect 25:1473–8. https://doi.org/10.1016/j.cmi.2019.04.020.

13. O’Neill LAJ, Netea MG. 11 May 2020. BCG-induced trained immunity: can it offer protection against COVID-19? Nat Rev Immunol. https://doi.org/10.1038/s41577-020-y.

14. Zwerling A, Behr MA, Verma A, Brewer TF, Menzies D, Pai M. 2019. The BCG world atlas: a database of global BCG vaccination policies and practices. PLoS Med 8:e1001012. https://doi.org/10.1371/journal.pmed.1001012.

15. Papke LE, Wooldridge JM. 1996. Econometric methods for fractional response variables with an application to 401(k) plan participation rates. J Appl Econom 11:619–32. https://doi.org/10.1002/(SICI)1099-1255(199611)11:6<619::AID-JAE418>3.0.CO;2-1

16. Papke LE, Wooldridge JM. 2008. Panel data methods for fractional response variables with an application to test pass rates. J Econom 145:121–33. https://doi.org/10.1016/jjeconom.2008.05.009.

17. Borjas GJ. 2020. Demographic determinants of testing incidence and Covid-19 infections in New York City neighborhoods. National Bureau of Economic Research Working Paper Series 26952. https://doi.org/10.3386/w26952.

18. Kissler SM, Tedijanto C, Goldstein E, Grad YH, Lipsitch M. 14 April 2020. Projecting the transmission dynamics of SARS-CoV-2 through the postpandemic period. Science. https://doi.org/10.1126/science.abb5793.

19. The Lancet. 4 April 2020. Redefining vulnerability in the era of COVID-19. Lancet 395. https://www.thelancetcom/action/showPdf?pii=S0140-6736%2820%2930757-1.

